# Association between HIV/AIDS, Medical Male Circumcision and Traditional Circumcision in Mozambique, 2015

**DOI:** 10.1101/2023.11.22.23298896

**Authors:** Hélio Inácio E. Militão, José Carlos Langa, Judite Monteiro Braga, Erika Valeska Rossetto, Cynthia Sema Baltazar, Timothy A. Kellogg

## Abstract

**Background:** The WHO AFRO region continues to be severely impacted by HIV and a global public health problem. In Mozambique, as of 2015, HIV prevalence was estimated to be 12.5% among adults. Medical male circumcision (MMC) has been promoted as a national prevention strategy to reduce the risk of HIV in men. We aimed to analyze the association between HIV infection, medical male circumcision and traditional male circumcision among men in Mozambique.

**Methods:** Cross-sectional data from the 2015 Mozambican National and Demographic Health Survey of Immunization, Malaria, and HIV/AIDS (IMASIDA) were used in this analysis. In this study, considered medical male circumcision (MMC) and medical circumcision (MC). Participants were asked about their circumcision status and where the circumcision was performed. In addition, blood samples were collected from participants and tested for HIV antibodies. All analyses were weighted and adjusted for the complex survey design to ensure results and approximate population parameters of interest. Chi-square tests and multiple logistic regression were used in the analyses to assess the associations between male circumcision and HIV infection.

**Results:** A total unweighted sample of 4733 men aged 15-49 consented to the survey and were interviewed. Of those who participated, 4236 consented to an HIV test. Nationally, 62.6% (95% CI 59.6-65.6) of men aged 15-49 years were circumcised. Traditional circumcision was the most common form of circumcision at 32.9% (95% CI 30.1-35.8), followed by MMC at 21.7% (95% CI 19.6-24.1), 8.0% (95% CI 6.5-9.9) did not know whether it was traditional or medical. The prevalence of HIV was highest at 13.4% among uncircumcised men (13.4%, 95% CI 11.3-15.7), and significantly lower among traditionally circumcised men (8.5%, 95% CI 6.8-10.6) and among medically circumcised men (7.5%, 95% CI 5.7-9.6). In multivariable analysis, men circumcised by a medical practitioner had almost 50% reduced odds of infection with HIV compared to uncircumcised men (aOR=0.52; 95% [CI=0.34-0.78], p=0.002), whereas men who were circumcised by traditional methods had a 29% reduced risk, but not significantly different than uncircumcised men (aOR=0.71; 95% [CI=0.47-1.07], p=0.098).

**Conclusion:** We found that HIV prevalence was lower among men aged 15-49 years who were circumcised, and the odds of being HIV positive was about 50% lower among men who were circumcised by a medical practitioner, suggesting a reduced risk of HIV infection. Although Mozambique has not achieved the UNAIDS goal of 80% men circumcised at the time of the IMASIDA 2015 survey, we encourage the continued expansion of voluntary medical male circumcision.

## Background

In 2018, over 37.9 million people were living with HIV worldwide, with 1.7 million people becoming newly infected with HIV [2, 3]. In Mozambique, as of 2015, approximately 1.8 million people were infected with HIV, with 120,000 new infections per year (UNAIDS 2021). Several randomized trials in Africa have provided evidence that MMC reduces the risk of female-to-male transmission of HIV by 60%[3]. Moreover, recent meta-analyses support the protective benefits of MMC in preventing HIV infection [4,5]. As a result, the World Health Organization (WHO) and the United Nations Programme on HIV/AIDS (UNAIDS) recommend that countries with generalized epidemics with low circumcision rates in the population introduce a strategy of providing MMC to uncircumcised men [9–13]. The World Health Organization (WHO) and the Joint United Nations Programme on HIV/AIDS (UNAIDS) identified 14 priority countries, including Mozambique, for MMC campaigns to reduce transmission of HIV. They set a target of 80% coverage for men aged 15-49 years [4,6,7] by 2018. The 2009 INSIDA, the last national HIV survey, found that 51% of males 15-49 years were circumcised. Most circumcised men were from four provinces. Niassa, Cabo Delgado, Nampula, and Inhambane -where most men undergo traditional circumcision (INSIDA 2009). In 2012, the Mozambique National Ministry of Health adopted a strategic plan focusing scale-up of MMC to seven provinces of the country - Maputo City, Maputo province, Gaza, Zambezia, Manica, Tete, and Sofala - that have low coverage of male circumcision. The plan set a target of two million males aged 10-49 years to be medically circumcised by 2017 to reduce HIV incidence [8].

In Mozambique, data from population-based surveys have contributed to estimating the prevalence of male circumcision among men, which assists in assessing if the target coverage of VMMC is being met. [9]. However, more information is necessary for determining the relative preventive benefits of traditional male circumcision in addition to MMC in preventing HIV infection. Using data from the 2015 IMASIDA survey, this study investigates the coverage of circumcision and the association between HIV infection and circumcision, both medical and traditional circumcision in Mozambique.

## Methods

### Survey Setting and Sample Description

The 2015 HIV/AIDS Indicator Survey in Mozambique - The National Indicators of Immunization, Malaria and HIV/AIDS Survey (IMASIDA 2015), was a nationally representative cross-sectional household survey conducted among children 6-59 months and adults aged 15-59 years using a standard AIS protocol. The Ministry of Health conducted it through the National Institute of Health, the National Public Health Directorate, and the National Statistics Institute with support from various collaborators and stakeholders in health. The methods used in IMASIDA 2015 have been previously described in detail[10]. Briefly, a stratified 2-stage cluster sampling design was used to select households, and within households, eligible participants were invited to participate. All habitual residents or visitors who spent the night before the interview in the selected households were eligible for the individual interview and blood sampling for those who consented. A trained health worker conducted a face-to-face interview with consenting participants in a private place inside or near the home. Data collection occurred between June and September 2015. In addition to the interview, informed consent was obtained from participants to draw blood for centralized HIV testing for survey purposes. The survey team offered rapid HIV testing using the prevailing national HIV testing algorithm to participants who wanted to know their HIV status during the survey visit.

### Measures

The questionnaire asked male participants for their age, marital status, educational level, residential setting, and employment status at the time of the interview. Interviewers also asked about their sexual behaviors, including the number of lifetime and recent sexual partners, condom use during the last time they had sex, self-reported symptoms of an STI, HIV knowledge, attitudes, perceived risk for HIV infection, HIV testing history, and, if tested previously, the result of their last HIV test. Comprehensive knowledge of HIV/AIDS was assessed using a standard set of knowledge questions previously used by other AIS and Demographic Health Surveys, exploring knowledge of transmission, prevention, treatment, and misconceptions related to HIV [10]. In this analysis, we focused on self-reported circumcision (the removal of the foreskin) status among men, when it occurred, the location (in their home, a health center, during an initiation rite), and who performed the circumcision (a traditional practitioner or health professional)[11]. A relative wealth index was generated for each household based on a detailed survey of the household’s ownership of selected assets, such as televisions and bicycles, livestock, materials used for housing construction, and types of water access and sanitation facilities, based on other demographic and health survey guidelines. This scale was categorized into five wealth quintiles and used to evaluate the association between household wealth and circumcision status[12]. Interviewers asked participants their opinion about the protective benefits of circumcision in preventing HIV infection, and if circumcised, if they believed men need to use condoms during sex.

### Specimen Collection and HIV testing

For male participants who consented to HIV testing, blood was drawn with a finger stick and collected on a filter paper using dried blood spots. Couriers transported the specimens to the central public health laboratory and tested for HIV using a sequential algorithm. Specimens were first analyzed using Vironostika HIV Ag/Ab (Biomerieux, France). Non-reactive specimens were considered negative for HIV. Reactive specimens were further tested by Murex HIV Ag/Ab Combination (DiaSorin, UK). Regardless of the Murex result, all reactive samples by Vironostika were further analyzed with a confirmatory test (Geenius ™ HIV 1/2 Rapid Confirmation Test, Bio-Rad, France). Samples with a negative PCR result were considered negative for HIV, and samples with a positive PCR result were considered positive for HIV [10].

### Statistical Analyses

All population-based percentages, odds ratios, and confidence intervals reported in this study were weighted so that estimates derived from this survey can approximate population parameters of interest. Statistical analyses were performed in Stata version 16.1 (Stata Corp, College Station, Texas), using the svyset command to account for the sampling design specifications (stratification, sample weighting, and clustering). To compare with other AIS surveys conducted elsewhere, we restricted the analysis to male participants aged 15 to 49. Weighted percentages and 95% confidence intervals (CIs) were generated for population-level characteristics to estimate circumcision status and the type of circumcision (medical circumcision or traditional) based on interview responses. To control for confounding, this analysis used multivariable logistic regression analysis to identify variables associated with circumcision. Since the analysis included the HIV results of the survey as a predictor variable, we limited the logistic regression analysis to men who consented to a blood draw for HIV testing.

### Ethical Considerations

The protocol for conducting the 2015 IMASIDA was approved by the ICF’s Internal Review Board, the Mozambique National Committee on Bioethics for Health, and the Division of Global HIV/AIDS at the U.S. Centers for Disease Control and Prevention. Ethical approval to conduct the IMASIDA 2015 survey was obtained from the National Committee for Bioethics in Health of Mozambique (Comité Nacional de Bioética para Saúde, CNBS) number: 127/CNBS/2013, the Institutional Review Board (IRB) of ICF International, and the US Centers for Disease Control and Prevention (CDC).

## Results

A total unweighted sample of 4773 men aged 15–49 years consented to the survey and were interviewed. Of those who participated, 4236 consented to an HIV test. Nationally, 62.6% (95% CI 59.6-65.6) of men aged 15-49 were circumcised. traditional circumcision was the most common form of circumcision at 32.9% (95% CI 30.1-35.8), followed by MMC at 21.7% (95% CI 19.6-24.1), with 8.0% (95% CI 6.5-9.9) circumcised but did not know the practitioner (Table 1). Men who received MMC tended to be younger, with 15-19 year-olds, the youngest age group, with the highest percentage of MMC at 33.1% (95% CI 29.4-37.1), whereas men 45-49 years, the oldest, had the highest percentage of receiving traditional circumcision at 43.5% (95% CI 37.1-50.0). In addition, the percentage of men who underwent medical circumcision was highest in urban areas at 35.9% (95% CI 31.7-40.4), whereas traditional circumcision was highest in rural settings at 38.9% (95% CI 35.1-42.9). Provinces with the highest percentages of any form of male circumcision were Niassa at 95.0% (95% CI 89.7-97.7), Cabo Delgado at 84.0% (95% CI 72.6-91.2), Nampula at 92.7% (95% CI (88.5-95.4), and Inhambane at 89.8% (95% CI 80.4-95.0), provinces where traditional circumcision is common. The provinces with the lowest levels of male circumcision include Tete at 9.2% (95% CI 5.6-14.8), Manica at 20.4% (95% CI 15.2-26.8), and Sofala at 20.1% (95% CI 14.6-27.1), provinces that are targeted for MMC scale-up (Table 1).

**Table 1.** Estimates of Male Circumcision by demographic characteristics of Mozambican Men, 15-49 years, IMASIDA, 2015.

| Characteristics | Uncircumcised<br>Weighted %<br>[95% CI] | Circumcised<br>Weighted %<br>[95% CI] | Type of Circumcision<br>Weighted % [95% CI] |  |  |
| --- | --- | --- | --- | --- | --- |
|  |  |  | Medical | Traditional | Don't Know<br>Practitioner |
| Total | 37.4 [34.4-40.4] | 62.6 [59.6-65.6] | 21.7 [19.6-24.1] | 32.9 [30.1-35.8] | 8.0 [6.5-9.9] |
| Age Group |  |  |  |  |  |
| 15-19 | 31.8 [28.0-35.8] | 68.2 [64.2-72.0] | 33.1 [29.4-37.1] | 27.7 [23.9-31.8] | 7.4 [5.7-9.6] |
| 20-24 | 36.7 [32.4-41.3] | 63.3 [58.7-67.6] | 23.8 [20.4-27.5] | 30.2 [26.6-34.1] | 9.2 [6.5-13.0] |
| 25-29 | 39.7 [35.0-44.6] | 60.3 [55.4-65.0] | 22.9 [19.3-26.9] | 29.1 [24.2-34.5] | 8.3 [6.1-11.1] |
| 30-34 | 41.1 [36.1-46.3] | 58.9 [53.7-63.9] | 15.7 [12.4-19.6] | 33.4 [29.1-38.0] | 9.9 [6.7-14.4] |
| 35-39 | 39.8 [33.6-46.2] | 60.2 [53.8-66.4] | 15.3 [11.9-19.4] | 38.8 [33.1-44.9] | 6.1 [4.1-9.1] |
| 40-44 | 39.5 [33.7-45.6] | 60.5 [54.4-66.3] | 12.1 [8.5-17.0] | 39.7 [34.1-45.7] | 8.6 [5.7-12.8] |
| 45-49 | 38 [32.0-44.5] | 62.0 [55.5-68.0] | 13.1 [9.6-17.5] | 43.5 [37.1-50.0] | 5.4 [3.4-8.6] |
| Residential Setting |  |  |  |  |  |
| Urban | 31.1 [27.7-34.8] | 68.9 [65.2-72.3] | 35.9 [31.7-40.4] | 23.7 [20.1-27.7] | 9.3 [6.4-13.3] |
| Rural | 41.4 [37.2-45.8] | 68.9 [65.2-72.3] | 12.4 [10.4-14.7] | 38.9 [35.1-42.9] | 7.2 [5.7-9.1] |
| Region |  |  |  |  |  |
| North | 9.4 [6.6-13.1] | 90.6 [86.9-93.4] | 14.9 [11.7-18.7] | 62.5 [57.7-67.0] | 13.2 [10.0-17.4] |
| Central | 73.3 [68.1-78.0] | 26.7 [22.0-31.9] | 13.5 [11.0-16.4] | 11.7 [8.2-16.5] | 1.4 [0.9-2.2] |
| South | 31.8 [27.6-36.4] | 68.2 [63.6-72.4] | 41.8 [36.8-47.0] | 17.3 [13.6-21.8] | 9 [6.7-12.1] |
| Province |  |  |  |  |  |
| Niassa | 5.0 [2.3-10.3] | 95.0 [89.7-97.7] | 10.1 [6.1-16.3] | 83.8 [74.7-90.1] | 1.1 [0.3-3.8] |
| Cabo Delgado | 16.0 [8.8-27.4] | 84.0 [72.6-91.2] | 14.1 [9.8-19.8] | 66.7 [55.7-76.1] | 3.2 [1.6-6.3] |
| Nampula | 7.3 [4.6-11.5] | 92.7 [88.5-95.4] | 16.5 [11.7-22.6] | 55.2 [49.4-60.8] | 21 [15.7-27.5] |
| Zambézia | 52.4 [40.9-63.7] | 47.6 [36.3-59.1] | 15.2 [10.2-22.2] | 31.9 [22.1-43.6] | 0.4 [0.1-1.5] |
| Tete | 90.8 [85.2-94.4] | 9.2 [5.6-14.8] | 6.2 [3.6-10.5] | 2.3 [1.1-4.8] | 0.7 [0.2-3.0] |
| Manica | 79.6 [73.2-84.8] | 20.4 [15.2-26.8] | 15.2 [10.4-21.6] | 1.8 [0.8-4.2] | 3.4 [1.9-5.9] |
| Sofala | 79.9 [72.9-85.4] | 20.1 [14.6-27.1] | 15.5 [11.3-21.0] | 3 [1.7-5.3] | 1.6 [0.7-3.9] |
| Inhambane | 10.2 [5.0-19.6] | 89.8 [80.4-95.0] | 31.1 [18.5-47.2] | 36 [23.9-50.1] | 22.8 [14.5-33.9] |
| Gaza | 52.5 [41.9-62.9] | 47.5 [37.1-58.1] | 41.1 [31.4-51.6] | 4.1 [2.1-8.0] | 2.2 [1.0-4.9] |
| Maputo Provincia | 31.4 [25.1-38.6] | 68.6 [61.4-74.9] | 41 [31.2-51.5] | 17.3 [11.1-26.0] | 10.2 [6.9-15.0] |
| Maputo Cidade | 28.9 [23.5-35.1] | 71.1 [64.9-76.5] | 50.6 [45.7-55.6] | 16.1 [12.1-21.1] | 4.3 [2.0-9.2] |
| Education Level |  |  |  |  |  |
| No education | 37.5 [29.3-46.5] | 62.5 [53.5-70.7] | 7.1 [4.2-11.9] | 49.5 [40.8-58.3] | 5.8 [3.5-9.7] |
| Primary | 42.2 [38.6-46.0] | 57.8 [54.0-61.4] | 13.6 [11.8-15.5] | 37.0 [33.6-40.7] | 7.1 [5.5-9.2] |
| Secondary | 31.5 [27.8-35.5] | 68.5 [78.4-90.2] | 34.9 [31.0-39.1] | 23.1 [20.0-26.5] | 10.5 [7.9-13.8] |
| Post-secondary | 14.8 [9.8-21.6] | 85.2 [59.6-65.6] | 69.8 [62.2-76.5] | 9.9 [6.0-15.9] | 5.5 [2.9-10.2] |
| Marital Status |  |  |  |  |  |
| Never married | 33.9 [30.3-37.6] | 66.1 [62.4-69.7] | 34.2 [30.6-38.0] | 24.5 [21.2-28.3] | 7.4 [5.6-9.8] |
| Married/living | 39.1 [35.6-42.7] | 60.9 [57.3-65.5] | 15.2 [13.2-17.5] | 37.3 [34.1-40.7] | 8.3 [6.6-10.5] |
| together |  |  |  |  |  |
| Widowed/divorced/<br>separated | 38.9 [31.5-46.9] | 61.1 [53.1-68.5] | 19.5 [15.2-24.8] | 32.7 [25.4-40.9] | 8.8 [5.3-14.4] |
| Religion |  |  |  |  |  |
| Christian | 36.4 [32.8-40.3] | 63.6 [59.7-67.2] | 26.2 [23.4-29.2] | 30.6 [26.9-34.5] | 6.7 [5.3-8.5] |
| Islam | 6.7 [4.6-9.5] | 93.3 [90.5-95.4] | 17.1 [13.1-21.9] | 62.3 [56.4-67.8] | 14.0 [8.5-22.0] |
| Traditional | 63.1 [55.0-70.4] | 36.9 [29.6-45.0] | 15.3 [10.7-21.2] | 13.9 [9.7-19.4] | 7.8 [4.9-12.0] |
| Other or no religion | 69.1 [64.1-73.6] | 30.9 [26.4-35.9] | 13.9 [11.2-17.1] | 12.2 [8.9-16.6] | 4.8 [3.2-7.1] |
| Wealth Index |  |  |  |  |  |
| Poorest | 34.4 [31.8-42.3] | 65.6 [59.0-71.6] | 7.5 [5.1-10.9] | 50.7 [44.1-57.3] | 7.4 [4.8-11.1] |
| Poorer | 36.9 [31.8-42.3] | 63.1 [57.7-68.2] | 9.7 [7.1-12.9] | 47.3 [42.0-52.7] | 6.2 [4.1-9.1] |
| Middle | 50.2 [42.9-57.6] | 49.8 [42.4-57.1] | 10.1 [7.7-13.3] | 31.9 [26.2-38.2] | 7.8 [5.1-11.6] |
| Richer | 38.3 [32.6-44.4] | 61.7 [55.6-67.4] | 24.3 [20.2-28.9] | 27.2 [22.6-32.3] | 10.2 [7.0-14.7] |
| Richest | 29.9 [26.3-33.8] | 70.1 [66.2-73.7] | 45.5 [41.4-49.8] | 16.1 [13.1-19.6] | 8.5 [6.3-11.3] |

The percentage of men with medical circumcision increased with higher education attainment, with the lowest percentage of MMC among men with no education at 7.1% (95% CI 4.2-11.9) and the highest for men with post-secondary education at 69.8% (95% CI 62.2-76.5). In contrast, traditional circumcision is highest for men without formal education at 49.5% (95% CI 40.8-58.3). Medical circumcision is highest for never married men at 34.2% (95% CI 30.6-38.0) and lowest for men currently married or in a union at 15.2% (95% CI 13.2-17.5). Men of Islamic faith have the highest percentage of male circumcision at 93.3% (95% CI 90.5-95.4), primarily from traditional circumcision, whereas men with no religious affiliation had the lowest percentage of circumcision at 30.9% (95% CI 26.4-35.9). The majority of men in the highest wealth level were circumcised at 70.1% (95% CI 66.2-73.7), with 45.5% (95% CI 41.4-49.8) medically circumcised compared to 7.5% (95% CI 5.1-10.9) in the lowest wealth category medically circumcised (Table 1).

### HIV Prevalence by circumcision status and other characteristics

Overall, 10.1% (95% CI 8.9-11.4) of men 15-49 years were positive for HIV (Table 2). Prevalence was highest among uncircumcised men at 13.4% (95% CI 11.3-15.7). Lower HIV prevalence was found among both traditional (8.5% [95% CI 6.8-10.6]) and medically circumcised (7.5% [95% CI 5.7-9.6]) (Table 2). Prevalence was highest 35–39-year-olds (17.5 95% CI 14.0-21.7); men living in urban areas (12.3% 95% CI 10.3-14.7); who are widowed (28.6% 95% CI 22.2-36.0); and among men living in Maputo Province (15.8% 95% CI 10.9-22.5.0). For sexual risk behaviors, HIV prevalence was 16.2% (95% CI 12.5-20.6) among men who had 6-9 lifetime partners, and 9.2% (95% CI 4.9-16.7) who had 3+ partners in the last 12 months (Table 2).

**Table 2.**
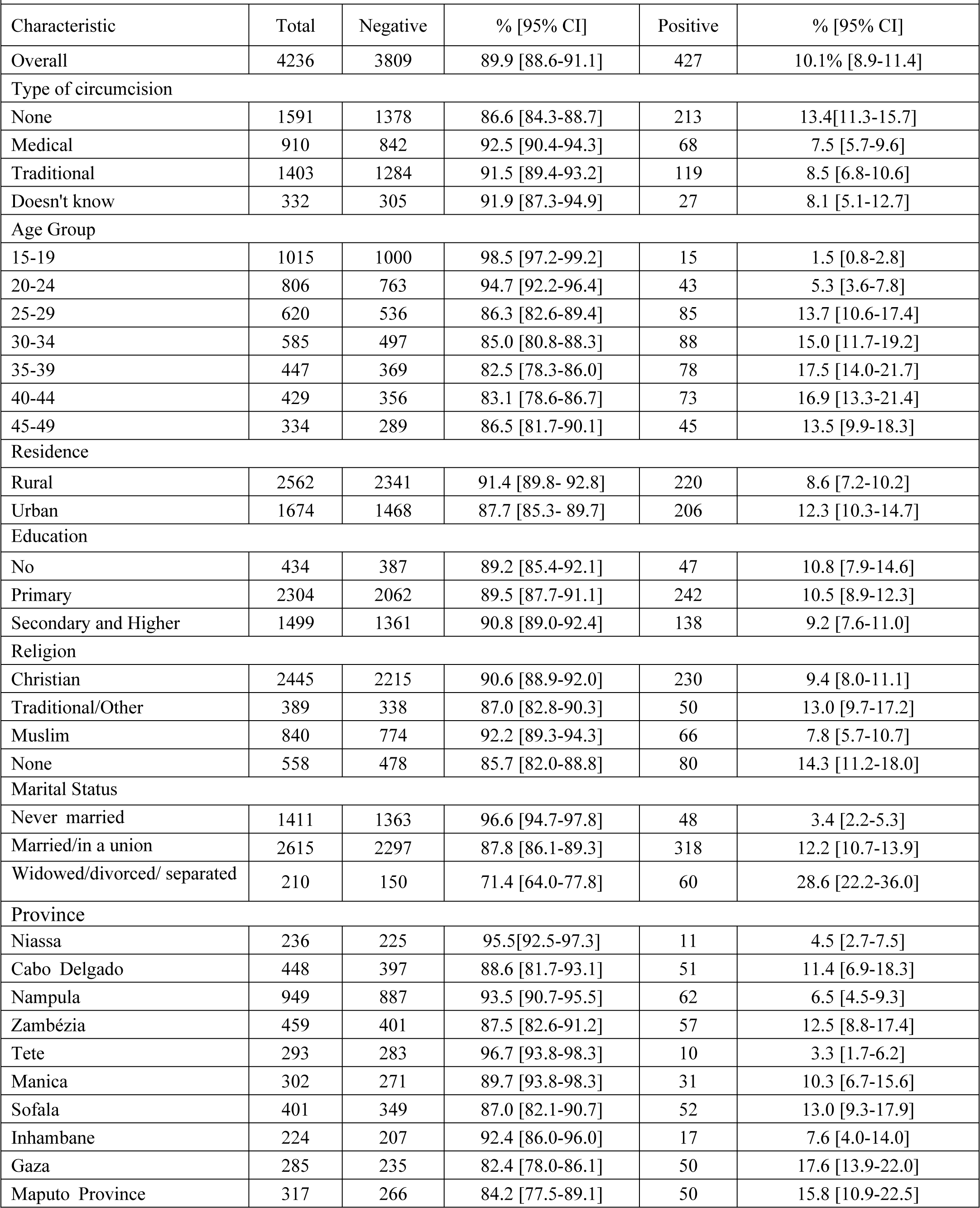
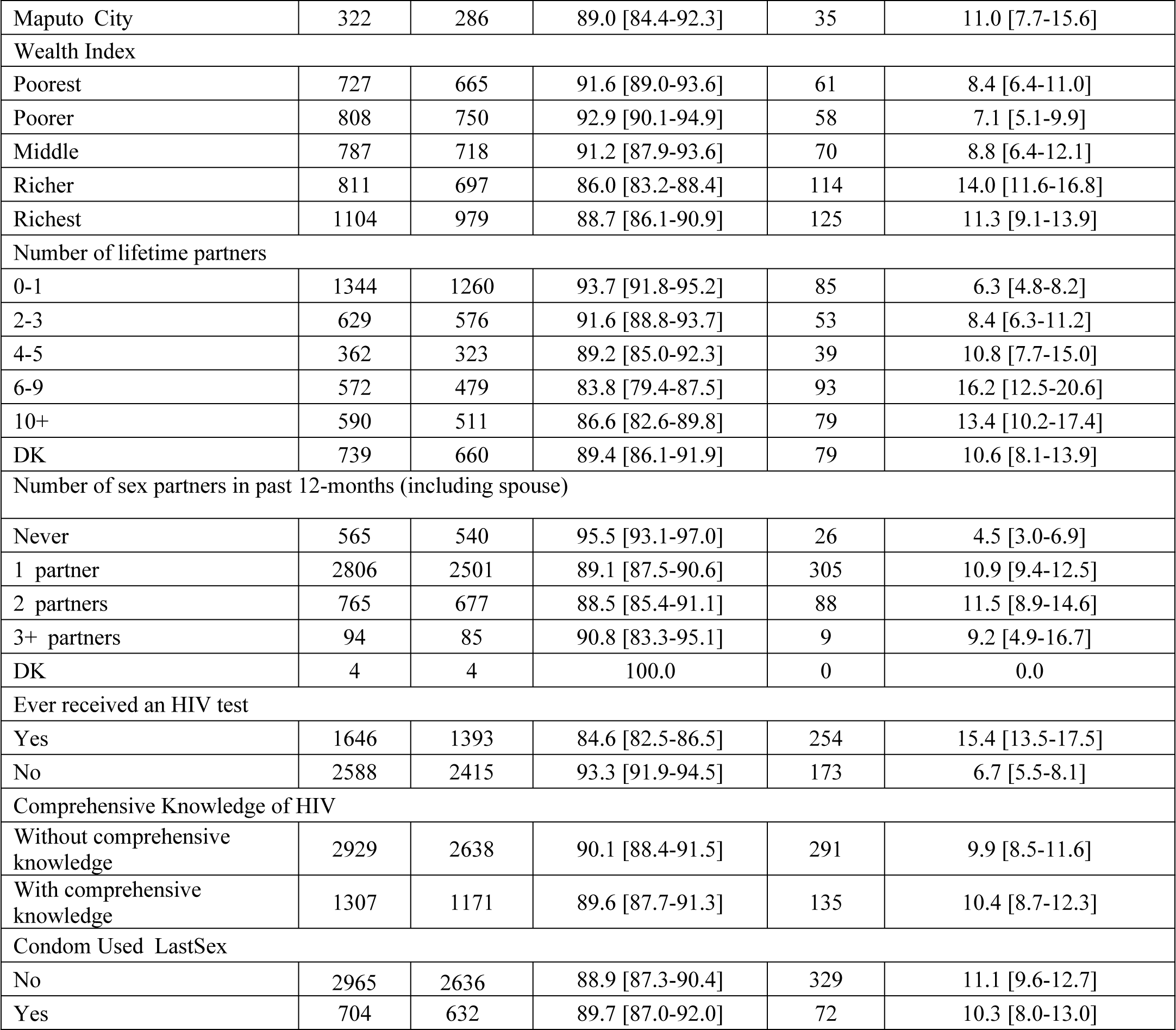
Estimates of HIV infection by socio-demographic characteristics, Mozambican Men, 15-49 years, IMASIDA, 2015.

| Characteristic | Total | Negative | % [95% CI] | Positive | % [95% CI] |
| --- | --- | --- | --- | --- | --- |
| Overall | 4236 | 3809 | 89.9 [88.6-91.1] | 427 | 10.1% [8.9-11.4] |
| Type of circumcision |  |  |  |  |  |
| None | 1591 | 1378 | 86.6 [84.3-88.7] | 213 | 13.4[11.3-15.7] |
| Medical | 910 | 842 | 92.5 [90.4-94.3] | 68 | 7.5 [5.7-9.6] |
| Traditional | 1403 | 1284 | 91.5 [89.4-93.2] | 119 | 8.5 [6.8-10.6] |
| Doesn't know | 332 | 305 | 91.9 [87.3-94.9] | 27 | 8.1 [5.1-12.7] |
| Age Group |  |  |  |  |  |
| 15-19 | 1015 | 1000 | 98.5 [97.2-99.2] | 15 | 1.5 [0.8-2.8] |
| 20-24 | 806 | 763 | 94.7 [92.2-96.4] | 43 | 5.3 [3.6-7.8] |
| 25-29 | 620 | 536 | 86.3 [82.6-89.4] | 85 | 13.7 [10.6-17.4] |
| 30-34 | 585 | 497 | 85.0 [80.8-88.3] | 88 | 15.0 [11.7-19.2] |
| 35-39 | 447 | 369 | 82.5 [78.3-86.0] | 78 | 17.5 [14.0-21.7] |
| 40-44 | 429 | 356 | 83.1 [78.6-86.7] | 73 | 16.9 [13.3-21.4] |
| 45-49 | 334 | 289 | 86.5 [81.7-90.1] | 45 | 13.5 [9.9-18.3] |
| Residence |  |  |  |  |  |
| Rural | 2562 | 2341 | 91.4 [89.8- 92.8] | 220 | 8.6 [7.2-10.2] |
| Urban | 1674 | 1468 | 87.7 [85.3- 89.7] | 206 | 12.3 [10.3-14.7] |
| Education |  |  |  |  |  |
| No | 434 | 387 | 89.2 [85.4-92.1] | 47 | 10.8 [7.9-14.6] |
| Primary | 2304 | 2062 | 89.5 [87.7-91.1] | 242 | 10.5 [8.9-12.3] |
| Secondary and Higher | 1499 | 1361 | 90.8 [89.0-92.4] | 138 | 9.2 [7.6-11.0] |
| Religion |  |  |  |  |  |
| Christian | 2445 | 2215 | 90.6 [88.9-92.0] | 230 | 9.4 [8.0-11.1] |
| Traditional/Other | 389 | 338 | 87.0 [82.8-90.3] | 50 | 13.0 [9.7-17.2] |
| Muslim | 840 | 774 | 92.2 [89.3-94.3] | 66 | 7.8 [5.7-10.7] |
| None | 558 | 478 | 85.7 [82.0-88.8] | 80 | 14.3 [11.2-18.0] |
| Marital Status |  |  |  |  |  |
| Never married | 1411 | 1363 | 96.6 [94.7-97.8] | 48 | 3.4 [2.2-5.3] |
| Married/in a union | 2615 | 2297 | 87.8 [86.1-89.3] | 318 | 12.2 [10.7-13.9] |
| Widowed/divorced/ separated | 210 | 150 | 71.4 [64.0-77.8] | 60 | 28.6 [22.2-36.0] |
| Province |  |  |  |  |  |
| Niassa | 236 | 225 | 95.5[92.5-97.3] | 11 | 4.5 [2.7-7.5] |
| Cabo Delgado | 448 | 397 | 88.6 [81.7-93.1] | 51 | 11.4 [6.9-18.3] |
| Nampula | 949 | 887 | 93.5 [90.7-95.5] | 62 | 6.5 [4.5-9.3] |
| Zambézia | 459 | 401 | 87.5 [82.6-91.2] | 57 | 12.5 [8.8-17.4] |
| Tete | 293 | 283 | 96.7 [93.8-98.3] | 10 | 3.3 [1.7-6.2] |
| Manica | 302 | 271 | 89.7 [93.8-98.3] | 31 | 10.3 [6.7-15.6] |
| Sofala | 401 | 349 | 87.0 [82.1-90.7] | 52 | 13.0 [9.3-17.9] |
| Inhambane | 224 | 207 | 92.4 [86.0-96.0] | 17 | 7.6 [4.0-14.0] |
| Gaza | 285 | 235 | 82.4 [78.0-86.1] | 50 | 17.6 [13.9-22.0] |
| Maputo Province | 317 | 266 | 84.2 [77.5-89.1] | 50 | 15.8 [10.9-22.5] |
| Maputo City | 322 | 286 | 89.0 [84.4-92.3] | 35 | 11.0 [7.7-15.6] |
| Wealth Index |  |  |  |  |  |
| Poorest | 727 | 665 | 91.6 [89.0-93.6] | 61 | 8.4 [6.4-11.0] |
| Poorer | 808 | 750 | 92.9 [90.1-94.9] | 58 | 7.1 [5.1-9.9] |
| Middle | 787 | 718 | 91.2 [87.9-93.6] | 70 | 8.8 [6.4-12.1] |
| Richer | 811 | 697 | 86.0 [83.2-88.4] | 114 | 14.0 [11.6-16.8] |
| Richest | 1104 | 979 | 88.7 [86.1-90.9] | 125 | 11.3 [9.1-13.9] |
| Number of lifetime partners |  |  |  |  |  |
| 0-1 | 1344 | 1260 | 93.7 [91.8-95.2] | 85 | 6.3 [4.8-8.2] |
| 2-3 | 629 | 576 | 91.6 [88.8-93.7] | 53 | 8.4 [6.3-11.2] |
| 4-5 | 362 | 323 | 89.2 [85.0-92.3] | 39 | 10.8 [7.7-15.0] |
| 6-9 | 572 | 479 | 83.8 [79.4-87.5] | 93 | 16.2 [12.5-20.6] |
| 10+ | 590 | 511 | 86.6 [82.6-89.8] | 79 | 13.4 [10.2-17.4] |
| DK | 739 | 660 | 89.4 [86.1-91.9] | 79 | 10.6 [8.1-13.9] |
| Number of sex partners in past 12-months (including spouse) |  |  |  |  |  |
| Never | 565 | 540 | 95.5 [93.1-97.0] | 26 | 4.5 [3.0-6.9] |
| 1 partner | 2806 | 2501 | 89.1 [87.5-90.6] | 305 | 10.9 [9.4-12.5] |
| 2 partners | 765 | 677 | 88.5 [85.4-91.1] | 88 | 11.5 [8.9-14.6] |
| 3+ partners | 94 | 85 | 90.8 [83.3-95.1] | 9 | 9.2 [4.9-16.7] |
| DK | 4 | 4 | 100.0 | 0 | 0.0 |
| Ever received an HIV test |  |  |  |  |  |
| Yes | 1646 | 1393 | 84.6 [82.5-86.5] | 254 | 15.4 [13.5-17.5] |
| No | 2588 | 2415 | 93.3 [91.9-94.5] | 173 | 6.7 [5.5-8.1] |
| Comprehensive Knowledge of HIV |  |  |  |  |  |
| Without comprehensive knowledge | 2929 | 2638 | 90.1 [88.4-91.5] | 291 | 9.9 [8.5-11.6] |
| With comprehensive knowledge | 1307 | 1171 | 89.6 [87.7-91.3] | 135 | 10.4 [8.7-12.3] |
| Condom Used LastSex |  |  |  |  |  |
| No | 2965 | 2636 | 88.9 [87.3-90.4] | 329 | 11.1 [9.6-12.7] |
| Yes | 704 | 632 | 89.7 [87.0-92.0] | 72 | 10.3 [8.0-13.0] |

### Factors Associated with HIV including Male Circumcision

Results from the multivariable analysis used to assess HIV infection and circumcision status are shown in Table 3. After controlling for demographic and sexual risk behaviors, medical circumcision was found to be significantly protective against HIV infection, with men circumcised by a medical practitioner at almost 50% lower risk for HIV (aOR 0.52 [95% CI 0.34-0.78]) compared with uncircumcised men. Traditional circumcision lessened the risk of HIV by about 30% (aOR 0.71 [0.47-1.07]) compared with uncircumcised men, but this association was not statistically significant (p=0.098). Increasing age was highly associated with HIV, as was living in an urban setting (aOR 1.55 [95% 1.01-1.42]), being widowed (aOR 2.46 [95% 1.14-5.31]) and having an STI in the last 12-months (aOR 2.22 [95% CI 1.54-3.20]).

**Table 3.** Multiple logistic regression analysis of the association between HIV and male circumcision and other predictors-in Mozambique-IMASIDA-2015.

| Characteristic | Unadjusted<br>OR (95% CI) | p-value | Adjusted<br>aOR (95% CI) | p- value |
| --- | --- | --- | --- | --- |
| **Type of circumcision** |  |  |  |  |
| Uncircumcised | Ref | -- | Ref | -- |
| Medical | 0.52 (0.38-0.72) | <0.001 | 0.52 (0.34-0.78) | 0.002 |
| Tradicional | 0.60 (0.45-0.82) | 0.001 | 0.71 (0.47-1.07) | 0.098 |
| Don't Know | 0.57 (0.34-0.97) | 0.036 | 0.22 (0.31-1.31) | 0.222 |
| **Age-Group** |  |  |  |  |
| 15-19 | Ref | -- | Ref | -- |
| 20-24 | 3.79 (1.9-7.7) | <0.001 | 1.86 (0.79-4.37) | 0.156 |
| 25-29 | 10.6 (5.2-21.8) | <0.001 | 4.21 (1.53-11.60) | 0.006 |
| 30-34 | 11.9 (6.1-23.3) | <0.001 | 5.55 (2.13-14.48) | <0.001 |
| 35-39 | 14.3 (6.9-29.3) | <0.001 | 6.32 (2.17-18.36) | 0.001 |
| 40-44 | 13.7 (6.7-27.9) | <0.001 | 6.55 (2.31-18.55) | <0.001 |
| 45-49 | 10.5 (5.0-22.0) | <0.001 | 4.83 (1.65-14.12) | 0.004 |
| **Residential Setting** |  |  |  |  |
| Rural | Ref |  | Ref | -- |
| Urban | 1.5 (1.1-1.9) | 0.004 | 1.55 (1.01-1.42) | 0.046 |
| **Education Level** |  |  |  |  |
| None | Ref | -- | Ref | -- |
| Primary | 0.96 (0.67-1.40) | 0.847 | 0.88 (0.56-1.42) | 0.613 |
| Secondary and Higher | 0.83 (0.56-1.23) | 0.564 | 0.79 (0.46-1.36) | 0.393 |
| **Religion** |  |  |  |  |
| Christian | Ref | -- | Ref | -- |
| Traditional/Other | 1.43 (0.99-20.7) | 0.052 | 1.24 (0.84-1.85) | 0.277 |
| Muslim | 0.82 (0.55-1.2) | 0.318 | 0.88 (0.53-1.47) | 0.626 |
| No Religion | 1.60 (1.1-2.2) | 0.003 | 1.66 (1.2-2.3) | 0.003 |
| **Marital Status** |  |  |  |  |
| Never Married | Ref | -- | Ref | -- |
| Married/Living Together | 3.91 (2.46-6.21) | <0.001 | 1.19 (0.54-1.47) | 0.633 |
| Widowed/Divorced/Separated | 11.29 (6.41-19.88) | <0.001 | 2.46 (1.14-5.31) | 0.003 |
| **Province** |  |  |  |  |
| Niassa | Ref |  | Ref | -- |
| Cabo Delgado | 2.71 (1.25-5.88) |  | 2.44 (1.14-5.18) | 0.020 |
| Nampula | 1.47 (0.76-2.85) |  | 1.31 (0.65-2.66) | 0.451 |
| Zambezia | 3.01 (1.54-5.87) |  | 1.88 (0.86-3.96) | 0.095 |
| Tete | 0.72 (0.31-1.68) |  | 0.34 (0.14-0.87) | 0.024 |
| Manica | 2.42 (1.18-4.96) |  | 1.10 (0.16-2.68) | 0.823 |
| Sofala | 3.16 (1.64-4.96) |  | 1.77 (0.80-3.93) | 0.160 |
| Inhambane | 1.74 (0.73-4.14) |  | 1.44 (0.54-3.83) | 0.462 |
| Gaza | 4.51 (2.46-8.27) |  | 2.83 (1.28-6.27) | 0.010 |
| Maputo Provincia | 3.97 (1.99-7.92) |  | 2.19 (0.97-4.98) | 0.060 |
| Maputo Cidade | 2.61 (1.33-5.11) |  | 1.44 (0.65-3.21) | 0.367 |
| Wealth Index |  |  |  |  |
| Poorest | Ref | -- | Ref | -- |
| Poorer | 0.84 (0.54-1.30) |  | 0.67 (0.40-1.12) | 0.130 |
| Middle | 1.05 (0.69-1.61) |  | 0.96 (0.58-1.60) | 0.887 |
| Richer | 1.77 (1.26-2.50) |  | 1.41 (0.87-2.28) | 0.166 |
| Richest | 1.38 (0.95-2.00) |  | 1.01 (0.53-1.93) | 0.975 |
| Lifetime sex partners |  |  |  |  |
| 0-1 | Ref |  | Ref | -- |
| 2-3 | 1.37 (0.92-2.04) |  | 0.93 (0.59-1.47) | 0.757 |
| 4-5 | 1.81 (1.16-2.82) |  | 1.01 (0.59-1.73) | 0.966 |
| 6-9 | 2.88 (1.89-4.38) |  | 1.57 (0.99-2.48) | 0.055 |
| 10+ | 2.30 (1.56-3.39) |  | 0.97 (0.65-1.45) | 0.893 |
| DK | 1.77 (1.17-2.67) |  | 0.98 (0.62-1.54) | 0.926 |
| Sex Partners last 12-months |  |  |  |  |
| No partners | Ref | -- | Ref | -- |
| 1 partner | 2.56 (1.65-3.97) | <0.001 | 0.78 (0.44-1.37) | 0.383 |
| 2 partners | 2.71 (1.61-4.59) | <0.001 | 0.72 (0.37-1.41) | 0.342 |
| 3+ partners | 2.13 (0.91-5.01) | 0.082 | 0.60 (0.21-1.73) | 0.346 |
| Ever been tested for HIV |  |  |  |  |
| Yes | Ref | -- | Ref | -- |
| No | 0.39 (0.31-0.49) | <0.001 | 0.52 (0.38-0.71) | <0.001 |
| Had any STI in last 12-months |  |  |  |  |
| No | Ref | -- | Ref | -- |
| Yes | 2.61 (1.84-3.71) | <0.001 | 2.22 (1.54-3.20) | <0.001 |
| With comprehensive knowledge about HIV |  |  |  |  |
| Yes | Ref | -- | Ref | -- |
| No | 1.04 (0.82-1.33) | <0.001 | 0.88 (0.68-1.49) | 0.375 |
| Condom use last sex |  |  |  |  |
| No | Ref | -- | Ref | -- |
| Yes | 0.92 (0.68-1.24) | 0.573 | 1.01 (0.68-1.49) | 0.979 |

## Discussion

In this nationally representative survey conducted in 2015, we found that Mozambican men 15-49 years who were medically circumcised were less likely to be infected with HIV by almost 50% compared to uncircumcised men after controlling for sexual behaviors, sociodemographic characteristics and other confounders, supporting evidence from other studies that MMC can be an effective method of HIV prevention. The protective benefit of traditional circumcision against HIV was less clear from this analysis as the risk for HIV infection, although 30% lower than uncircumcised men, was not significantly different from that for uncircumcised men. A second finding of this study found that nationally, 62.6% of all men between 15-49 years have been circumcised of any type, well below the goal of 80% by 2020 and 90% by 2025. These findings have several important implications for improving national programs that encourage MMC. Traditional circumcision remains the more common form of circumcision among men in Mozambique as of 2015. However, MMC is becoming slightly more common among men of younger age. In the 2009 INSIDA survey, the previous national survey, MMC accounted for only 37% of circumcisions among men between 15-24 years who were circumcised (INSIDA 2009). In the 2015 IMASIDA survey, MC accounted for 44% of all circumcised men between 15-24 years, an increase of 7%. However, when analyzing 15-24-year-olds within the provinces that were targeted for MMC scale-up (Maputo City, Maputo province, Gaza, Zambezia, Manica, Tete, and Sofala), the percentage of MMC among all circumcised men in this age-group was 76%. Targeting specific age groups for MMC may help achieve the national MC goal. A recent modeling study that assessed targeting certain age groups for MMC programs in Mozambique found that between 2009 through the end of September 2017, MMC programs increased coverage of MC from 27% to 48% for 10-49 years-olds, primarily among 10 to 29 year-olds, and projected to avert over 67,000 new infections by 2030. The authors concluded that maintaining MMC in this age group offers a cost-effective strategy [8].

Mozambique continues to face a generalized HIV epidemic, with an estimated prevalence of 10.1% among men 15-49 years in 2015 and 9.0% in 2021[10,13]. Despite HIV prevention interventions over the past three decades, HIV and AIDS remain a significant public health challenge in Mozambique [4,11,13–15].

Few studies deal with the association between HIV and Medical Circumcision and traditional circumcision in Mozambique, even though several studies have been conducted in Africa and worldwide related to evidence and knowledge of HIV prevention through MC and traditional circumcision.

We found that the most frequently circumcised age group was 15-19, followed by 20-24 year olds. Most MC was performed in urban areas compared to traditional circumcision. This may be associated with the fact that men in urban areas have more access to available information about the risks of traditional circumcision. Men who did not receive either MC or traditional circumcision had a higher prevalence of HIV compared to men who received circumcision.

Regarding the protection of circumcision against HIV / AIDS, it was found that the majority who responded that they knew the methods of protection against HIV / AIDS had a lower prevalence compared to those who did not know the methods of protection, contrary to the level of education that there was a higher prevalence in men of secondary and higher education, needing more analysis on this phenomenon. These data on protection from HIV/AIDS were also referenced in a study conducted in Uganda which shows that individuals with knowledge of HIV protection methods tend to have a lower prevalence compared to those who do not know, the difference is that this study conducted in Uganda covered fewer circumcised men because it included a smaller sample, and added that cultural issues, fear of pain or complication of circumcision, may explain the low level of circumcision [16–18].

### Limitations

The authors recognize several limitations to this analysis. First, self-reported responses conducted during surveys may be subject to memory and social desirability biases and thus may incorrectly measure behaviors. Although the definition of circumcision was provided during the interview, including a description of how circumcision is conducted, respondents might not be knowledgeable about specifics of the surgical procedure or recall the exact location of where it was performed or who performed it and may be unable to distinguish between the two types of circumcision. Also, cross-sectional surveys cannot assess causality, as we cannot ascertain whether a man’s positive status occurred before or after the traditional circumcision Since it was a study that used the IMASIDA for secondary analysis, in some cases there were some categories of missing values due to unanswered questions.

Despite these limitations, this study provides information to policymakers and governments about the benefits of MMC as strategy for reducing the burden of HIV among men in Mozambique, especially in the areas of the country where traditional circumcision is not performed routinely. Moreover, this analysis will help gauge progress towards meeting national goals of MC coverage among Mozambican men between 15-49 years.

### Conclusion

We conclude that male circumcision was protective against HIV infection in Mozambique. In 2015, almost 2 in 3 men aged 15-49 in Mozambique were circumcised. In this study it was found that the higher the age the higher the frequency of HIV. Traditional circumcision occurs more in adults than at younger ages and most of them are from rural areas.

Urban areas tend to practice more medical circumcision than traditional circumcision, and most men in these areas have a higher prevalence of HIV compared to rural areas where traditional circumcision is a common practice. HIV prevalence tends to be higher in the Catholic religion.

It was found that men who perform traditional circumcision are more likely to be HIV positive than those who practice voluntary male medical circumcision, and HIV prevalence was higher among uncircumcised men compared to circumcised men. In this study we found that uncircumcised men were more likely to have HIV infection than circumcised men. We found an inverse association between HIV infection and medical/traditional circumcision, table 3. We encourage adolescents, boys and men aged 15-29 to undergo MMC in rural areas, and sensitize uncircumcised adults to choose MMC over traditional MMC which has a higher risk for HIV transmission.

### What is already known on this topic

- Medical male circumcision (MMC) is one of the crucial strategies for the reduction of HIV/AIDS in the world and in Mozambique.
- Medical circumcision protects against HIV infection

### What this study adds

- Predominance of traditional circumcision compared to voluntary medical male circumcision in Mozambique.
- MC had a greater protective effect than traditional circumcision.
- Education is not associated with the test result in the multiple analysis.
- The higher the wealth quintile the higher the HIV prevalence in non-circumcised groups.

### Financing

The Mozambique Field Epidemiology and Laboratory Training Program (FELTP) was funded through the PEPFAR cooperative agreement NU2GGH002021.

## Data Availability

Availability of data The authors of this manuscript make all the data inherent in this manuscript available without restriction to all requesters as soon as necessary. The datasets for this study are available on “Demographic and Health Surveys” (DHS) website https://dhsprogram.com/data/dataset/Mozambique_Standard-AIS_2015.cfm?flag=0

## Authors Contributions

HM, TK, EVR were responsible for designing the study. HM was responsible for data analysis and interpretation and wrote the manuscript. JMB and JPL supervised these analyses. TK supports the revision. EVR and CSB supervised and approved of the entire process (protocol to manuscript). All authors read and approved the preliminary version of the manuscript.

## Acknowledgments

Ministry of Health, Moz-FELTP coordinators from the National Institute of Health (INS) and Dr Jahit Sacarlal, from Faculty of Medicine of Eduardo Mondlane University.

I also thank Joshua Fortmann, Epidemiology Fellow, CDC Mozambique for critical analysis, interpretation, and support in describing the results.

